# Change in the neutrophil-to-lymphocyte ratio after a 3-day orbital flight: A preregistered secondary analysis of NASA open data from the Inspiration4 civilian crew

**DOI:** 10.64898/2026.10.16.26365757

**Authors:** Víctor Junior Godínez-Vázquez

## Abstract

**Background:** Commercial spaceflight now carries civilians on short missions, yet their stress-axis physiology is poorly described. Earlier shuttle data suggest that sympathetic responses predominate after short flights and glucocorticoid responses after long ones. The neutrophil-to-lymphocyte ratio (NLR) is a low-cost readout of both.

**Methods:** Preregistered secondary analysis of open clinical laboratory data (NASA Open Science Data Repository OSD-569 and OSD-575) from the four Inspiration4 crew members, each sampled seven times: three draws before launch, one the day after return (R+1) and three during recovery (R+45 to R+194). Log-transformed NLR was modelled with a linear mixed model with a random intercept per crew member and Kenward-Roger inference. Lymphocyte and eosinophil counts and C-reactive protein were secondary outcomes with false discovery rate correction.

**Results:** All 28 draws were analysed. NLR at R+1 did not differ from pre-flight (fold change 1.04; 95% CI 0.81 to 1.34; p = 0.73); two crew members increased (+25% and +46%) and two decreased (−27% and−11%). During recovery NLR was 23% lower than pre-flight (fold change 0.77; 95% CI 0.64 to 0.92; p = 0.005). No secondary outcome changed after correction; lymphocytes were 18% lower at R+1 (95% CI −35% to +4%; q = 0.31). Between-person differences explained 58% of the variance. An ordinary regression ignoring repeated measures missed the recovery effect (p = 0.08).

**Conclusions:** A 3-day orbital flight produced no consistent leukocyte stress signature one day after return. The lower NLR during recovery suggests that pre-flight values, not recovery values, deviate from the resting state, which has implications for how baselines are defined in short-duration spaceflight studies.

## Introduction

Space has been framed as humanity’s new frontier since the first decade of the space program (1). As technologies for transport and energy storage beyond Earth advance, the life sciences must advance with them. Astrobiology, space bioprocess engineering and bioastronautics serve different purposes, but they share one context: living organisms, including the human body, operating in an off-world environment whose effects are still incompletely understood (2).

From a clinical perspective, spaceflight carries well-characterized hazards. Ionizing radiation and microgravity affect multiple organs and systems and raise the risk of cancer, infection and metabolic disease, while the closed, artificial atmosphere of a spacecraft adds risks that are less well defined (3). Microgravity in orbit is a state of continuous free fall: the spacecraft’s horizontal velocity keeps it falling around Earth, so occupants experience near-weightlessness. Experiments in plants have shown how this state alters gravitropic and phototropic responses, physiology and gene expression, with consequences for bioregenerative life support (4,5).

Space radiation consists mainly of atomic nuclei travelling close to the speed of light, stripped of their electrons. It comes from three sources: particles trapped in Earth’s magnetic field, particles ejected by the Sun, and galactic cosmic rays, which are high-energy protons and heavier nuclei from outside the solar system (6). All are ionizing: they carry enough energy to remove electrons from atoms and alter the matter they cross. On Earth, the atmosphere absorbs and reflects much of the Sun’s ionizing electromagnetic radiation, a protection that is largely absent in space (7). Even at low doses, ionizing radiation carries measurable cancer risk: a meta-analysis of 26 epidemiological studies published between 2006 and 2017, including about 91,000 solid cancers and 13,000 leukemias, found excess risks compatible with those observed in atomic bomb survivors (8).

Beyond these chronic hazards, launch and landing are acute physiological stressors. In shuttle astronauts, landing was followed by neutrophilia and lymphopenia, and the mechanism depended on mission duration: sympathetic responses predominated after short flights, whereas longer flights showed glucocorticoid-mediated changes attributed to cumulative deconditioning (9). Both pathways raise the neutrophil-to-lymphocyte ratio (NLR): catecholamines through acute neutrophil demargination, and glucocorticoids through demargination plus redistribution of lymphocytes out of the circulation. A fall in eosinophils is more specific to glucocorticoid action. NLR is a ratio of two fractions of the same white-cell count, so it is not distorted by post-flight shifts in plasma volume.

NLR is also a clinical biomarker in its own right. In the general population it has been associated with mortality (10), and over short follow-up intervals its changes can reflect disordered inflammation and immunity, because it summarizes the balance between innate immunity, carried mainly by neutrophils, and adaptive immunity, carried by lymphocytes. An imbalance can point to acute or chronic inflammation, and changes in NLR over time have been proposed as markers of immune dysregulation, disease severity and mortality, although their interpretation depends on the context in which they are measured (11).

Commercial spaceflight has changed who flies. Civilian crews with less selection and training now fly short missions, and the first openly available clinical data from such a crew come from the 3-day SpaceX Inspiration4 mission of September 2021 (12,13). Whether the stress-axis pattern described in career astronauts applies to civilians on very short flights is unknown.

We therefore asked whether NLR changes within each Inspiration4 crew member from the pre-flight level to the day after return and during recovery, and whether eosinophils indicate a glucocorticoid component. The hypotheses, outcomes, exclusion rules and analysis code were registered before the analysis (14).

## Methods

### Design and registration

This is a secondary analysis of existing open data with a single-group, within-subject repeated-measures design. The analysis plan, including hypotheses, outcomes, exclusion rules and code, was registered on the Open Science Framework before the confirmatory analysis (14). Quality control of the source files preceded registration and is disclosed there; the primary outcome was not summarized by phase before registration.

### Participants and data sources

The sample was the complete crew of the Inspiration4 mission (n = 4 civilian adults, C001–C004), who flew for 3 days in low Earth orbit in September 2021 (12,13). Each crew member had seven blood draws: L-92, L-44 and L-3 days before launch, and R+1, R+45, R+82 and R+194 days after return. No clinical blood draws were taken in flight. Complete blood count (whole blood) was obtained from NASA Open Science Data Repository study OSD-569 (15), and the comprehensive metabolic panel and cardiovascular multiplex panel (serum) from OSD-575 (16). Files were downloaded on 15 October 2026 and archived with SHA-256 checksums.

### Variables

Flight phase was the predictor, with three levels: pre-flight (L-92, L-44, L-3; reference), return (R+1) and recovery (R+45, R+82, R+194). The primary outcome was NLR, computed as neutrophils (%) divided by lymphocytes (%). Secondary outcomes were absolute lymphocyte and eosinophil counts (white blood cell count × percentage) and C-reactive protein (CRP). All outcomes were natural-log transformed.

### Quality control and exclusions

Four rules were fixed before analysis and applied uniformly:

1. Absolute differential counts that disagreed with white blood cell count × percentage by more than 15% were replaced by the recomputed value (one draw, C003 at L-92, where the published columns were shifted).
2. Cardiovascular-panel values were excluded at a timepoint whose median across crew was below one-third of the analyte’s overall median, indicating an assay batch effect (L-44; 4 CRP values).
3. Glucose below 55 mg/dL in asymptomatic adults, consistent with in-vitro glycolysis, led to exclusion of glucose and potassium at that draw (R+45).
4. Analytes with an identical value in three or more samples were treated as censored and not analysed (haptoglobin).

No observation of the primary outcome was excluded.

### Statistical analysis

Each outcome was modelled with a linear mixed model, fitted by restricted maximum likelihood, with phase as a fixed effect and a random intercept per crew member, in R 4.3.3 (17) with lme4 (18). Inference used Kenward-Roger small-sample degrees of freedom (19). Contrasts against pre-flight were back-transformed and reported as fold changes with 95% confidence intervals. The primary hypothesis was tested two-sided at α = 0.05. Marginal and conditional R^²^ (20) and the intraclass correlation for crew were reported. The return-versus-pre-flight contrasts of the three secondary outcomes were corrected with the Benjamini-Hochberg false discovery rate (21). As a design-based check, an exact sign-flip test was applied to the four individual changes; with four participants its minimum attainable p value is 0.125. Pre-specified sensitivity analysis refitted the CRP model including L-44. To show the effect of ignoring repeated measures, ordinary least squares regression was also fitted. The false-positive rate of the planned model was 5.2% in 1,000 simulated datasets with this design and no phase effect.

## Results

All 28 draws from the four crew members were analysed for the primary outcome. Median NLR was 1.79 (interquartile range 1.42–2.39) before flight, 2.03 (1.88–2.12) at R+1 and 1.50 (1.17–1.87) during recovery.

### Primary outcome

NLR at R+1 did not differ from pre-flight (Table 1; Figure 1B), so the primary hypothesis was not supported. During recovery, NLR was 23% lower than pre-flight, and the confidence interval excluded no change. The omnibus test for phase was F(2, 22) = 5.99, p = 0.008. Phase explained 16% of the variance (marginal R² = 0.16), and phase plus crew 65% (conditional R² = 0.65); the intraclass correlation for crew was 0.58. The model converged without singularity.

**Table 1.** Change in NLR relative to pre-flight (linear mixed model, Kenward-Roger df = 22)

| Contrast | Fold change | 95% CI | p |
| --- | --- | --- | --- |
| Return (R+1) vs pre-flight | 1.04 | 0.81 to 1.34 | 0.73 |
| Recovery (R+45 to R+194) vs pre-flight | 0.77 | 0.64 to 0.92 | 0.005 |

**Figure 1.**
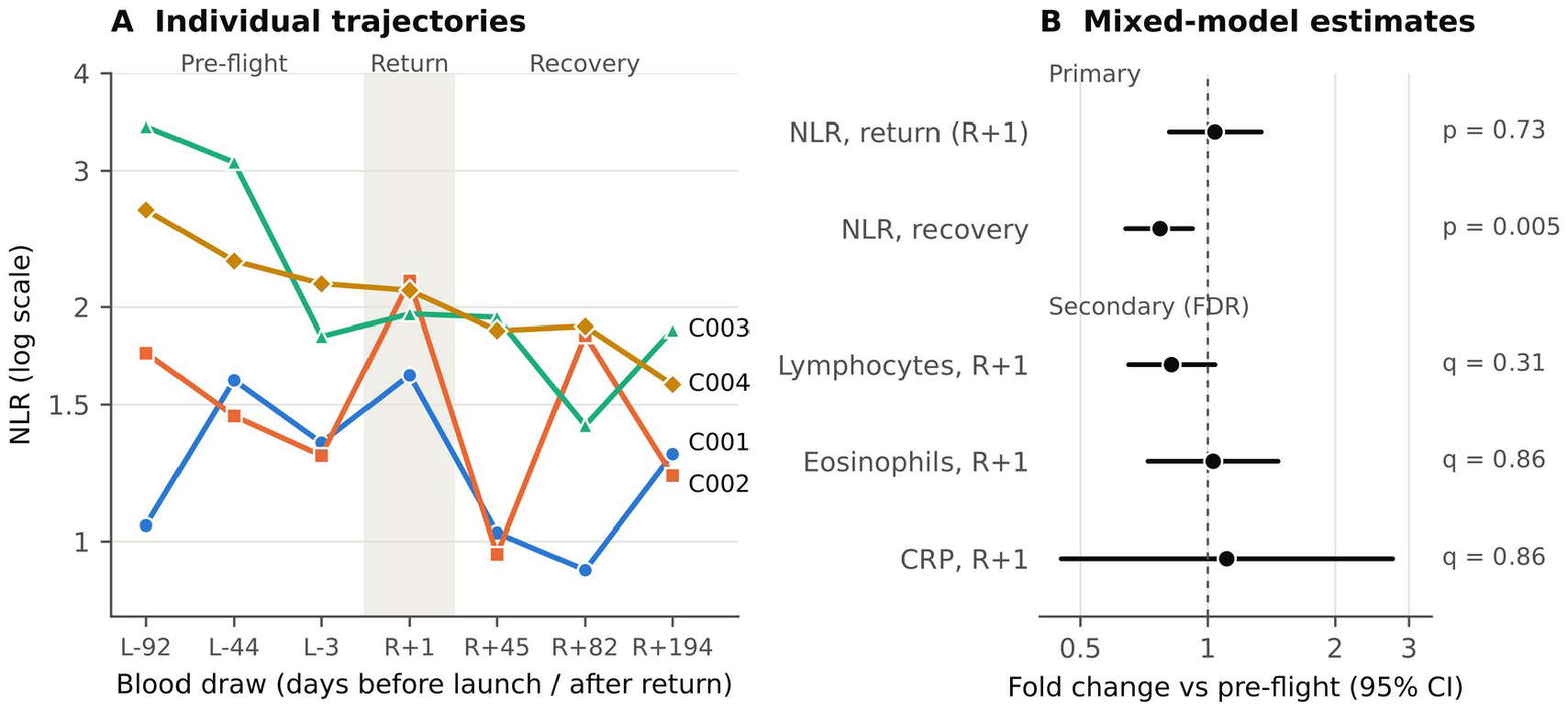
Neutrophil-to-lymphocyte ratio (NLR) in the four Inspiration4 crew members. (A) NLR at each of the seven blood draws, one line per crew member, on a log scale; the shaded band marks the day after return (R+1). (B) Fold change relative to pre-flight with 95% confidence intervals from linear mixed models with a random intercept per crew member and Kenward-Roger inference; values below 1 indicate a decrease. Secondary outcomes are shown for R+1 with Benjamini-Hochberg q values; C-reactive protein (CRP) excludes the L-44 batch.

### Individual responses

The direction of change at R+1 was inconsistent across the crew: NLR rose by 25% in C001 and 46% in C002, and fell by 27% in C003 and 11% in C004 relative to each person’s pre-flight mean. The exact sign-flip test gave p = 0.88. Two crew members (C003 and C004) had their highest NLR at the earliest pre-flight draws (L-92 and L-44), and their values declined towards launch (Table 2; Figure 1A).

**Table 2.** NLR by crew member and timepoint.

| Crew | L-92 | L-44 | L-3 | R+1 | R+45 | R+82 | R+194 |
| --- | --- | --- | --- | --- | --- | --- | --- |
| C001 | 1.05 | 1.61 | 1.34 | 1.64 | 1.03 | 0.92 | 1.29 |
| C002 | 1.75 | 1.45 | 1.29 | 2.16 | 0.96 | 1.84 | 1.21 |
| C003 | 3.42 | 3.08 | 1.84 | 1.96 | 1.94 | 1.41 | 1.87 |
| C004 | 2.67 | 2.29 | 2.14 | 2.10 | 1.87 | 1.89 | 1.59 |

### Secondary outcomes

No secondary outcome changed at R+1 after false discovery rate correction (Table 3). Lymphocyte count was 18% lower at R+1, in the hypothesized direction, but the interval included no change. Eosinophil count did not change, so the pre-specified rule for a glucocorticoid component was not met. The lymphocyte model was singular (crew variance estimated as zero).

**Table 3.** Secondary outcomes at R+1 relative to pre-flight.

| Outcome | n | Fold change | 95% CI | p | q (FDR) |
| --- | --- | --- | --- | --- | --- |
| Lymphocytes, absolute | 28 | 0.82 | 0.65 to 1.04 | 0.10 | 0.31 |
| Eosinophils, absolute | 28 | 1.03 | 0.72 to 1.47 | 0.86 | 0.86 |
| C-reactive protein* | 24 | 1.11 | 0.45 to 2.74 | 0.82 | 0.86 |

\*L-44 excluded (assay batch effect); not blind to timepoint medians.

### Sensitivity and model comparison

Including the L-44 CRP values changed the R+1 fold change from 1.11 to 2.11 (95% CI 0.68 to 6.59), showing the influence of the batch effect. Ordinary least squares regression that ignored repeated measures gave larger standard errors and did not detect the recovery effect (p = 0.08 versus 0.005 in the mixed model).

## Discussion

In the first civilian crew with openly released clinical data, a 3-day orbital flight did not produce a consistent rise in NLR one day after return. Two crew members increased and two decreased, and neither lymphocytes nor eosinophils changed beyond chance. The clearest signal was unexpected: NLR during recovery was about a quarter lower than before flight.

### The day after return

The absence of a consistent R+1 response fits the mechanism described by Stowe et al. (9). After short flights, the landing response in shuttle astronauts was predominantly sympathetic, and catecholamine-driven neutrophil demargination resolves within hours. A draw taken about one day after splashdown may therefore have missed it. A third possibility is that a response occurred and was fully compensated within the sampling interval; resting leukocyte measurements cannot distinguish a buffered response from an absent one. The unchanged eosinophil count argues against a glucocorticoid component, as expected when exposure is too short to cause the deconditioning that drives it after longer missions. Our data cannot separate a missed transient response, a compensated response and a true absence of response.

### Recovery lower than pre-flight

The lower NLR during recovery suggests that the pre-flight values, rather than the recovery values, departed from the resting state. The pattern does not resemble anticipatory stress before launch: in two crew members NLR was highest three months before launch (L-92) and declined towards L-3. Training load, travel, subclinical infection or other events in the pre-flight period are plausible explanations, but none can be tested with these data, and this interpretation is exploratory. The practical implication is methodological. A single pre-flight draw would have produced a different baseline for each crew member, and studies of short missions should collect several pre-flight samples and examine their stability before using them as the reference.

### Why the analysis model matters

Between-person differences accounted for 58% of the variance in NLR. An ordinary regression that treated the 28 draws as independent mixed this variation into the error term and failed to detect the recovery effect, which the mixed model identified. With few participants and many repeated draws, which is the typical structure of crewed spaceflight data, analysing within-person change is not optional.

### Open data quality

The source files contained a shifted row of absolute counts, glucose values consistent with delayed serum separation, an assay batch effect at one timepoint and a censored analyte. Each was detectable by simple consistency checks and was handled by a rule fixed before analysis. These findings illustrate both the value of open data and the need to check it before reuse; they will be reported to the repository maintainers.

### Limitations

The sample is four people, and the exact sign-flip test cannot reach conventional significance with four participants. There is no concurrent ground control group, so changes at return cannot be attributed to spaceflight alone. The crew comprised four healthy adults, so the analysis cannot address whether responses differ by factors that modify lipoprotein and glucocorticoid physiology, such as adiposity, sex, exogenous sex-steroid use, which can lower HDL cholesterol by about half (22), or smoking history; this limitation will become more consequential as commercial crews are selected less stringently than career astronauts. No blood was drawn in flight or within hours of landing, and no cortisol or catecholamine measurements are openly available, so the mechanisms are inferred from leukocyte patterns. Sampling windows were fixed by the original study, the laboratory reference intervals of one crew member changed after flight, and CRP was not analysed blind to timepoint medians.

### Future directions

This study tests only the first, measurable link of a longer chain, and its null result may reflect what was measured rather than an absent effect. Adrenal glucocorticoid synthesis depends on cholesterol delivered to mitochondria, where the side-chain cleavage reaction begins steroidogenesis (24). Under stress, the lipoprotein sources are not interchangeable: in mice, loss of scavenger receptor class B type I (SR-BI, encoded by *SCARB1*) abolished stress-induced glucocorticoid production even when low-density lipoprotein was raised about sixfold (27), and corticotropin upregulates *SCARB1* to increase cholesterol supply for steroidogenesis (28). Human loss-of-function states follow the same pattern: carriers of *SCARB1* variants and patients with LDL-receptor deficiency have resting cortisol within the reference range but a reduced response to corticotropin stimulation (26,29). Adrenal reserve is therefore invisible to resting measurement. SR-BI loss of function also raises HDL cholesterol and coronary risk (25), linking this axis to cardiovascular disease.

This matters for spaceflight because HDL cholesterol fell on landing day in shuttle crews (23), and because the glucocorticoid-mediated pattern described by Stowe et al. emerged only after longer missions (9). Both observations are compatible with a buffered system in which compensation is maintained over short exposures and lost with cumulative duration. Commercial missions could test this directly by pairing lipoprotein quantity and function with corticotropin-stimulated, rather than resting, glucocorticoid measurement, with dense sampling around landing and missions of differing duration.

### Conclusion

A 3-day orbital flight produced no consistent leukocyte stress signature one day after return in its civilian crew, while NLR during recovery was lower than before flight. Short-duration spaceflight studies need multiple pre-flight samples, sampling within hours of landing, analyses that model within-person change, and measurements taken under provocation rather than at rest. As space opens to civilians, these open, preregistered and reproducible methods are part of extending that frontier safely.

## Data Availability

All data used in this study are openly available from the NASA Open Science Data Repository: OSD-569 (doi:10.26030/e0rx-sh52) and OSD-575 (doi:10.26030/mc5d-p710). All data produced in the present work (the derived analysis file, the quality-control log, the analysis code and the output tables) are available online at Zenodo (doi:10.5281/zenodo.XXXXXXX). The preregistered analysis plan is available at the Open Science Framework (doi:10.17605/OSF.IO/W3NP2).

https://osdr.nasa.gov/bio/repo/data/studies/OSD-569

https://osdr.nasa.gov/bio/repo/data/studies/OSD-575

https://doi.org/10.17605/OSF.IO/W3NP2

## Declarations

### Data availability

The source data are openly available from the NASA Open Science Data Repository: OSD-569 (doi:10.26030/e0rx-sh52) and OSD-575 (doi:10.26030/mc5d-p710). The derived analysis file, quality-control log and analysis code are available at the Open Science Framework project linked to the preregistration (doi:10.17605/OSF.IO/W3NP2).

### Ethics

Secondary analysis of de-identified, publicly released data from participants who consented to open data sharing under the original investigators’ ethics approvals. No contact with participants. Ethics committee review was not required for this secondary analysis of openly available data.

### Author contributions

V.J.G.-V. conceived the study, designed and registered the analysis plan, interpreted the results and wrote the manuscript.

### Use of AI tools

An AI assistant (Claude, Anthropic) was used to support data quality control, code development and testing, and drafting of the manuscript. The author designed the study, verified all analyses and references, and takes full responsibility for the content.

## Acknowledgements

The author thanks the Inspiration4 crew for consenting to open data sharing, and the investigators and NASA Open Science Data Repository staff who collected, curated and released the data.

